# Cluster-Specific Urban Contexts Associated with High Levels of Sleep Impairment and Daytime Sleepiness: Findings from the Urbasan Collaborative Study

**DOI:** 10.1101/2024.12.10.24318755

**Authors:** Philippe Voruz, Marco Vieira Ruas, Noé Fellay, Noemi Romano, Michelangelo Mussini, Mathieu Saubade, Vincent Faivre, Vincent Gremeaux, Ophélia Jeanneret, Quentin Tonnerre, Patrick Beetschen, Marie-Noëlle Domon-Aubort, Dario Spini, Bengt Kayser, Daniel Rappo, Stéphane Joost

## Abstract

**Introduction:** Impaired sleep is a global health concern. However, the environmental factors contributing to sleep impairment in urban settings are still not well understood.

**Methodology:** This study involved 179 participants from a Swiss municipality (Yverdon-les-Bains), where sleep quality and diurnal sleepiness were measured using validated questionnaires, alongside environmental and geo-referenced data.

**Results:** The findings revealed a high prevalence of sleep disorders across diverse demographic groups (respectively 15.6% for diurnal sleepiness and 91.1% for significantly altered sleep quality). Additionally, sleep disorders were associated with both environmental and socio-demographic factors. Geospatial analysis identified clusters of sleep disturbances in specific neighborhoods, with distinct associations to specific sub-scores (factors) of the sleep evaluation.

**Conclusion:** Assessing sleep in urban environments is crucial, as it is linked to elevated levels of sleepiness. Environmental and socio-demographic variables play significant roles in these disturbances. The incorporation of geospatial analyses allows for a more precise identification of patterns within the city, offering opportunities for tailored interventions to address the different patterns of sleep disorders.

## 1. Introduction

Sleep disorders represent a critical public health issue, carrying not only clinical implications but also significant social and economic consequences [1]. These disorders are increasingly recognized not just as isolated conditions or symptoms but as factors intricately linked to other health conditions [2]. Moreover, they are now being identified as risk factors for various diseases, including those affecting the central nervous system, oncological health, and the cardiovascular system [2]. In this context, the prevention and early identification of sleep disorders is essential.

Research indicates that the living environment significantly impacts sleep quality [3]. Factors such as noise [4] and pollutants [5] have been linked to sleep disturbances. Consequently, the habitat and its surroundings present crucial opportunities for intervention. However, identifying at-risk areas and providing reliable data to enable effective action by responsible authorities requires large-scale collaborative studies conducted across diverse environments [6] [7]. These studies, often referred to as remote digital health studies, involve data collection through computers, smartphones, and wearable technology. Since 2016, such studies have become increasingly prevalent [8]. They offer several advantages, including targeting specific participant profiles, providing incentives or nudges, and reducing overall study complexity [8]. In the context of evaluating sleep quality, these experimental paradigms can help not only assess the prevalence of sleep disorders in the general population but also identify environmental and socio-economic factors influencing sleep quality, as well as their spatial distribution.

### This study aimed to achieve three objectives

#### Prevalence Assessment

To evaluate the prevalence of sleep disorders and sleepiness in a population-based sample from Yverdon-les-Bains, a medium-sized Swiss city with approximately 30,000 inhabitants in 2022 and a population density of 2,644 inhabitants per square kilometer. Switzerland, recognized for its high standard of living, provided a unique setting to test the hypothesis of observing significant levels of sleep impairment.

#### Relationship Analysis

Based on existing literature [4, 5] to examine the relationships between sleepiness and environmental as well as socio-economic factors, with the hypothesis that sleep quality and sleepiness would negatively correlate with these markers.

#### Spatial Analysis

Using validated statistical methodologies, to conduct exploratory spatial [9].

## 2. Methodology

### 2.1. General procedure of the Urbasan project

Urbasan is a collaborative online platform running on a dedicated website (https://yverdon.urbasan.ch/). It enables the progressive constitution of an e-cohort, within the framework of remote digital health studies [8]. The platform was developed in collaboration between the Institute of Environmental Engineering (IIE) at the Ecole Polytechnique Fédérale de Lausanne (EPFL), the Center for Primary Care and Public Health in Lausanne (Unisanté), the Swiss Centre of Expertise in Life Course Research (LIVES) at the University of Lausanne, the Lausanne University Hospital (CHUV), the City of Lausanne, the City of Yverdon-les-Bains and the Institut d’ingénierie des Médias (MEI) of the Haute Ecole d’Ingénierie et de Gestion du Canton de Vaud (HEIG-VD). This tool enabling continuous evaluation of the quality of physical and mental health using psychometrically validated questionnaires, is implemented in the town of Yverdon-les-Bains since 2022. The aim of the Urbasan e-cohort platform is to recruit 2’000 volunteer participants aged 18 and over who live in the commune of Yverdon-les-Bains by 2026. The following data are collected from participants: i) Individual data; ii) Socio-economic status; iii) Environmental characteristics of the place of residence; iv) Characteristics of the place of study or work; v) Physical activity; vi) General health data; vii) Sleep. To see detailed data inclusion within the Urbasan project, see Table 1.

**Table 1.** Data collected in the context of the Urbasan study.

| **Domain** | **Type of data collected** |
| --- | --- |
| Individual data | Surname, first name, e-mail address, home address (this information is used to georeference participants at their place of residence), gender, date of birth, marital status, nationality, mother tongue(s) (based on the questionnaires provided by the Swiss Household Panel, SHP; [10]) |
| Socio-economic status | Level of education, professional occupation (based on the questionnaires provided by the Swiss Household Panel, SHP; [10]) |
| Environmental characteristics of the place of residence | Walking distance to shops and facilities, level of vegetation, facilities encouraging or discouraging physical activity, safety aspects, road and rail noise as evaluated by the participants (based on specific analysis described in detail above) |
| Characteristics of the place of study or work | Distance from home, means of transport, infrastructure that encourages or discourages physical activity |
| Physical activity | Type, duration and intensity (based on the IPAQ-SF questionnaire [11]) |
| Health data | Height and weight, perception of personal health |
| Sleep | Sleep disorders (PSQI; [12]); daytime sleepiness (ESS; [13]) |

#### Ethics

The study was conducted in accordance with the Helsinki declarations and was approved by the ethics committee of the Faculty of Social and Political Sciences of the University of Lausanne (CER-SSP UNIL no C-SSP-112019-00002).

#### Data availability

All data produced in the present study are available upon reasonable request to the authors

**Figure 1.**
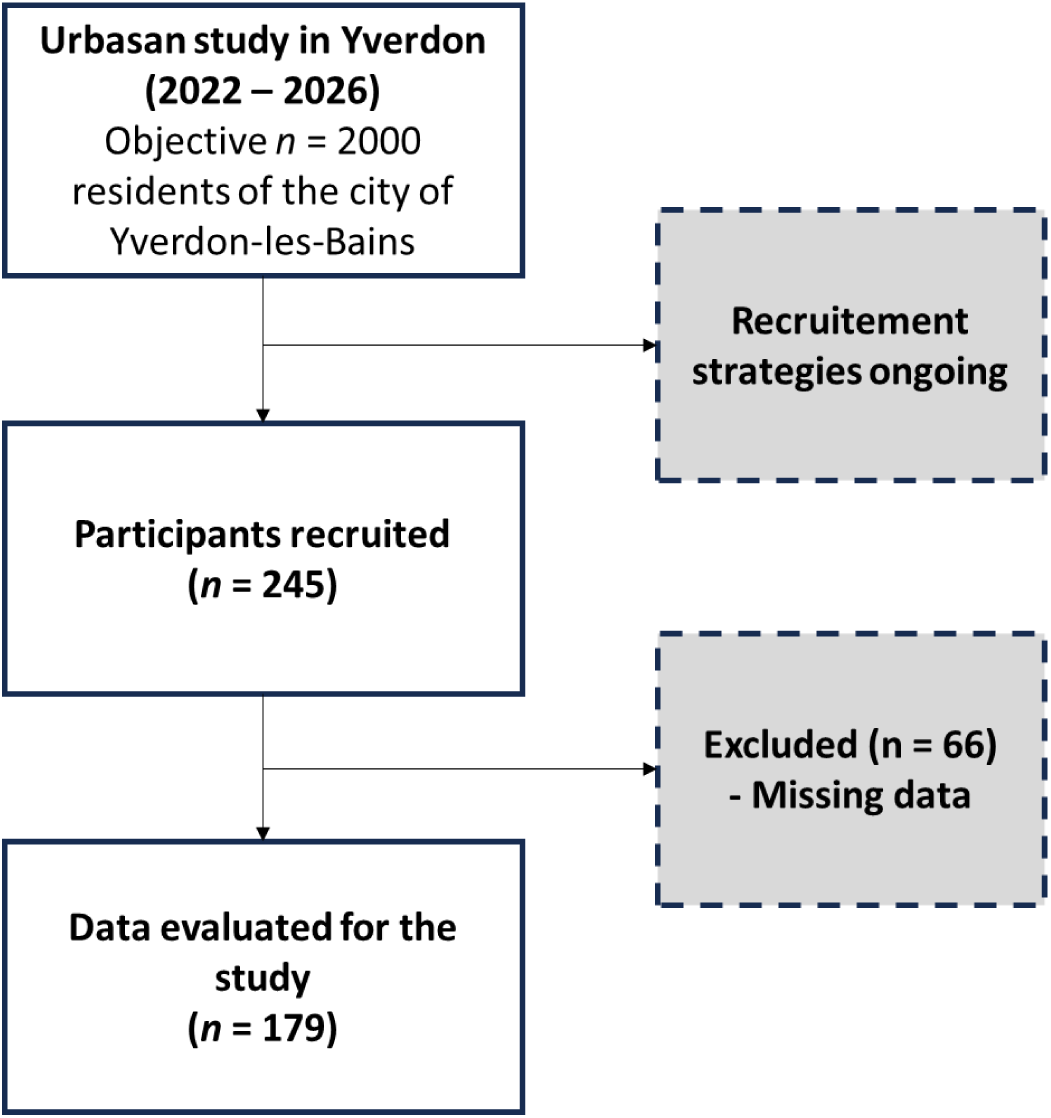
Flowchart of the Urbasan e-cohort in the city of Yverdon-les-Bains.

### 2.2. Population

In this pilot study, the objective was to evaluate an initial sample representing 10% of the expected final sample (n = 245/2000). This sample was recruited using communication strategies developed in collaboration with the town’s sports and urban planning department.

### 2.3. Data extraction for the study

In the context of this study, specific data have been extracted from the database.

#### 2.3.1. Sleep measures

*Epworth Sleepiness Scale (ESS; Johns [13]):* This validated questionnaire allows to identify individuals who may be suffering from excessive daytime sleepiness. Based on psychometric considerations, we determined a conservative clinical cut-off (>10), which suggests at least a moderate level of daytime sleepiness.

*Pittsburg Sleepiness Quality Index (PSQI; Buysse, Reynolds III [12]):* This validated questionnaire of sleep quality and disturbances over the past month. Factor analysis revealed explores seven different components: i) Subjective sleep quality; ii) Sleep latency; iii) Sleep duration; iv) Habitual sleep efficiency; v) Sleep disorders; vi) Use of sleeping pills; vii) Daytime dysfunction. Based on a recent systematic review that examined various clinical cut-off points, we chose a more conservative threshold (>10) rather than the more commonly used one (>5) [14]. Since this study is being conducted in a global context rather than a clinical setting, we decided on this conservative approach to more accurately capture significant changes in sleep patterns.

#### 2.3.2. Environmental and socio-economic variables

*The Normalized Difference Vegetation Index (NDVI)* makes it possible to assess vegetation density. It was computed on the basis of Sentinel2 satellite imagery (https://landsat.gsfc.nasa.gov/) from the United States Geological Survey (USGS). The index is calculated from spectrometric data with a spatial resolution of 30x30m at the red and near-infrared bands (NDVI = [Near Infrared - Red] / [Near Infrared + Red]. We used Google Earth Engine (GEE; https://earthengine.google.com/) to process 22 images recorded during the months of June, July and August 2023. We first created a single composite image by calculating the median value for each pixel across the range of images and then computed the NDVI. NDVI values range from [-1 to +1], where negative values represent water bodies, and high positive values indicate dense vegetation. Values near zero correspond to barren surfaces such as rock, asphalt, or sand [15].

*The Atmospherically Resistant Vegetation Index (ARVI)* was calculated on the same 22 images to assess vegetation density while minimizing the influence of atmospheric conditions, such as clouds, aerosols, and other particulate matter, which can interfere with the accuracy of satellite-based measurements [16].

*Land Surface Temperature (LST))* was computed on the basis of Landsat 8 & 9 satellite imagery (https://landsat.gsfc.nasa.gov/) from the United States Geological Survey (USGS) to get a proxy variable for potential urban heat islands. It was calculated on the basis of 21 images recorded between June and August 2023, to which we applied the statistical mono-window algorithm developed by Ermida, Soares [17].

*Road and railway noise:* to assess nocturnal traffic noise levels across Yverdon, this study made use of the sonBASE georeferenced database developed by the Swiss Federal Office for the Environment [18]. This database offers comprehensive information on nighttime road and rail noise exposure by means of a georeferenced 10 x 10 meters regular grid. The initial noise models were developed in 2008, based on extensive traffic data covering 72,000 kilometers of roads and 3,000 kilometers of rail. Here we used the most recent version of sonBASE (2015). Total noise exposure from road and rail traffic for each grid cell was calculated employing the formula from Goelzer, Hansen [19].

*Air Pollution:* the average concentrations of air pollutants per inhabited hectare were provided by Meteotest, under mandate from the OFEV [20]. This data includes nitrogen dioxide (NO_2_), as well as fine particulate matter smaller than 2.5 microns (PM2.5) and smaller than 10 microns (PM10) with a spatial resolution of 20 m.

*Risk Of Poverty Index (AROP):* the deprivation index used is a relative poverty indicator that expresses the percentage of people at risk of income poverty with the cut-off point set at 60% of median equivalised income [21]. The reference values used in this study are the yearly median income of private households for the canton of Vaud where the city Yverdon is located (CHF 80’280).

### 2.4. Statistical Analysis

#### 2.4.1. Statistics for behavioral and environmental data

Firstly, descriptive analyses (mean; standard deviation; range) are proposed for the entire cohort. Secondly, the prevalence of sleep disorders is analyzed using clinical cut-offs for the ESS and PSQI. Thirdly, because of the non-parametric data, Spearman correlations were performed between the sleep data (PSQI; ESS) and the environmental data, in order to assess the relationships between these variables.

#### 2.4.2. Statistics for spatial data

Using the geographical coordinates of participants’ place of residence, the Getis-Ord Gi* statistics [22, 23] was calculated with the GeoDa software (v.1.22; [24]). This statistic makes it possible to assess the spatial dependence and to identify geographic clusters of the variable of interest through the investigated territory. Specifically, the Getis-Ord Gi* compares the sum of individual’s PSQI values within a given neighborhood (spatial lag) to the sum of PSQI values across the entire study area. The statistic yields a Z-score, the corresponding null hypothesis assuming random spatial patterning of the values under analysis. Significance testing was conducted using a conditional randomization procedure with 999 permutations. Three classes are built based on this approach and displayed on maps: i) statistically significant positive Z-scores that indicate clustering of high PSQI values (hotspots); ii) statistically significant negative Z-scores that indicate clustering of low PSQI values (coldspots); iii) non-significant Z-scores that are neutral locations showing no spatial dependence.

## 3. Results

### 3.1. Demographics

Of the 245 participants, 179 provided complete data. The descriptive data from this sample reveal an average age of 48.2 years (± 14.2; [distribution: 18 - 79 years]), 60.4% of the sample were women and the average level of education was 3.91 (± 1.27). Average body mass index (BMI) was 24.28 (± 3.95). Based on the clinical categories defined by the World Health Organization, 63.10% of the sample had a ‘normal’ BMI, 0.02% an ‘underweight’ BMI, 26.20% of the sample ‘overweight’ and finally 0.90% of the sample was ‘obese’.

### 3.2. Prevalence of self-reported sleep disorders and daytime sleepiness

*Sleep (PSQI) and daytime sleepiness (ESS):* analysis revealed a considerable prevalence of sleep disorders. In particular, 15.6% of participants had a pathological score on the self-assessment of sleepiness (ESS: >10), while 91.1% had a pathological score on the self-assessment of sleep quality (PSQI: > 10) (see Fig. 2).

**Figure 2.**
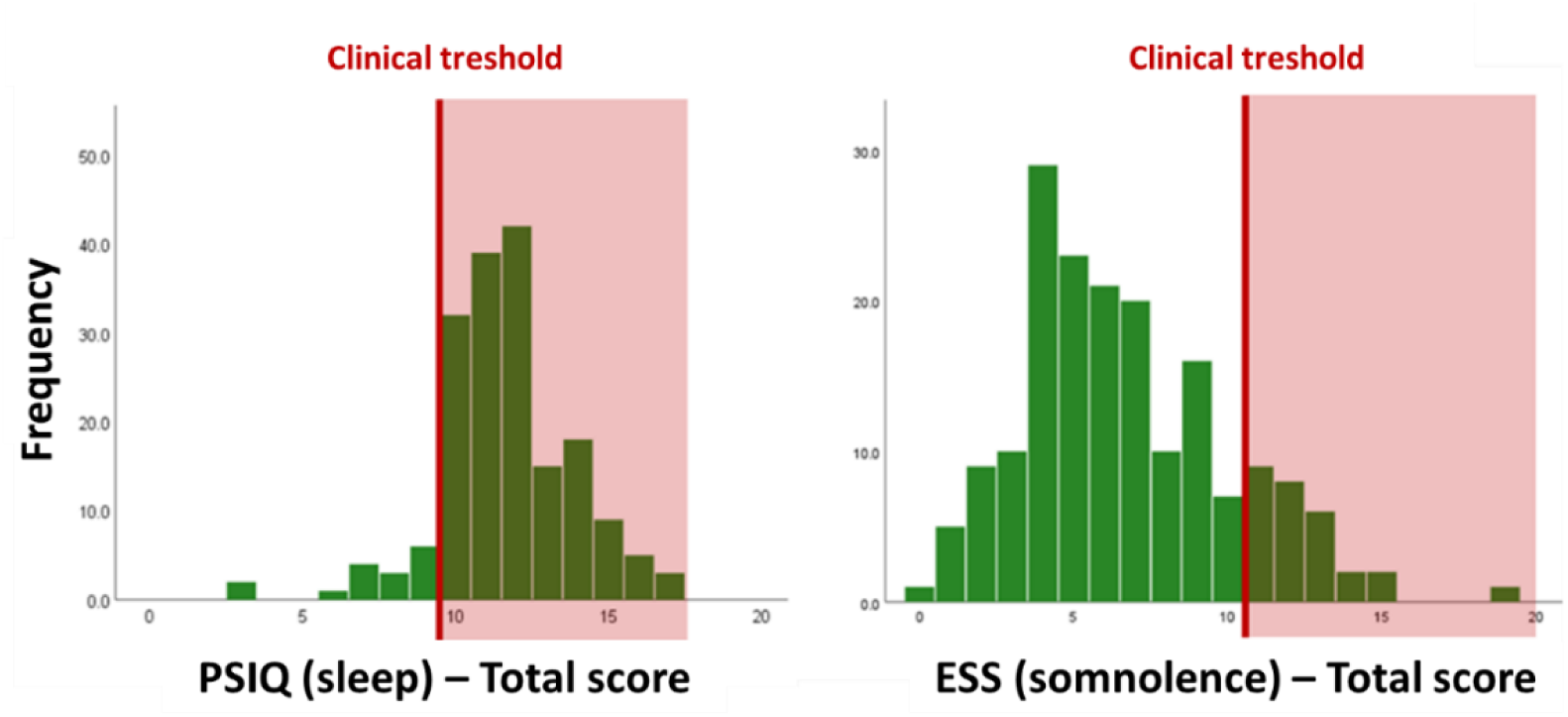
Frequency of scores obtained for the Pittsburgh Sleep Quality Index (PSQI) and the Epworth Sleepiness Scale (ESS), as well as the clinical cut-offs (in red), making it possible to delineate the pathological scores for the two self-questionnaires

### 3.3. Relationship between sleep and environmental variables

Spearman correlation analysis on the ESS score revealed significant associations with ARVI score (*r* = -.133; *p* = .038), NDVI score (*r* = -.126, *p* = .046) and the Rail Noise at Night (*r* = .134, *p* = .038). All other associations were non-significant (*p* > .061).

Spearman correlation analysis on the PSQI (total score) revealed significant association with the AROP (*r* = -.136, *p* = .035). Spearman correlation analysis on the subscores of the PSQI revealed significant relationships for specific results. The subscore PSQI – Subjective Sleep was associated to LST (*r* = -.136, *p* = .035), ARVI (*r* = .124, *p* = .049), NDVI (*r* = .141, *p* = .030), Road Noise at Day (*r* = .157, *p* = .018), Rail Noise at Night (*r* = -.153, *p* = .020), and AROP (*r* = .129, *p* = .043). The subscore PSQI – Medication was associated to Road Noise at Day (*r* = -.138, *p* = .033), Road Noise at Night (*r* = -.127, *p* = .046) and the AROP (*r* = -.219, *p* = .043). All other associations were non-significant (*p* > .119).

In order to check for the potential effects of confounding variables (sex; BMI; age) previously suggested in the literature [25], correlation analyses were performed on these confounding variables. In this context, the analyses did not reveal any significant associations between sleep scores and those socio-demographic variables (*p* > .071).

### 3.4. Spatial analysis results

The Getis-Ord analysis of the total PSQI score did not identify significant clusters (neither hotspots nor coldspots), suggesting a generally high mean PSQI randomly distributed across the entire city. However, a post-hoc Getis-Ord analysis of the significant subscores (Subjective Sleep, Medication, and Latency) revealed distinct clusters of both hotspots and coldspots within the city. Notably, we observed a clear coldspot cluster for the PSQI – Subjective Sleep subscore (circled in blue), while a similar cluster was found for the PSQI – Medication subscore, but with hotspots instead. This indicates that this specific neighborhood is associated with both higher perceived subjective sleep quality and increased medication use, suggesting a potential mediating role of sleep medication in shaping individual’s subjective sleep perceptions. Additionally, a hotspot cluster for the PSQI – Latency subscore was detected in the same neighborhood, indicating worse objective sleep evaluations (e.g., longer sleep latency), thereby highlighting a discrepancy between subjective sleep perception and objective sleep measures.

**Figure 2.**
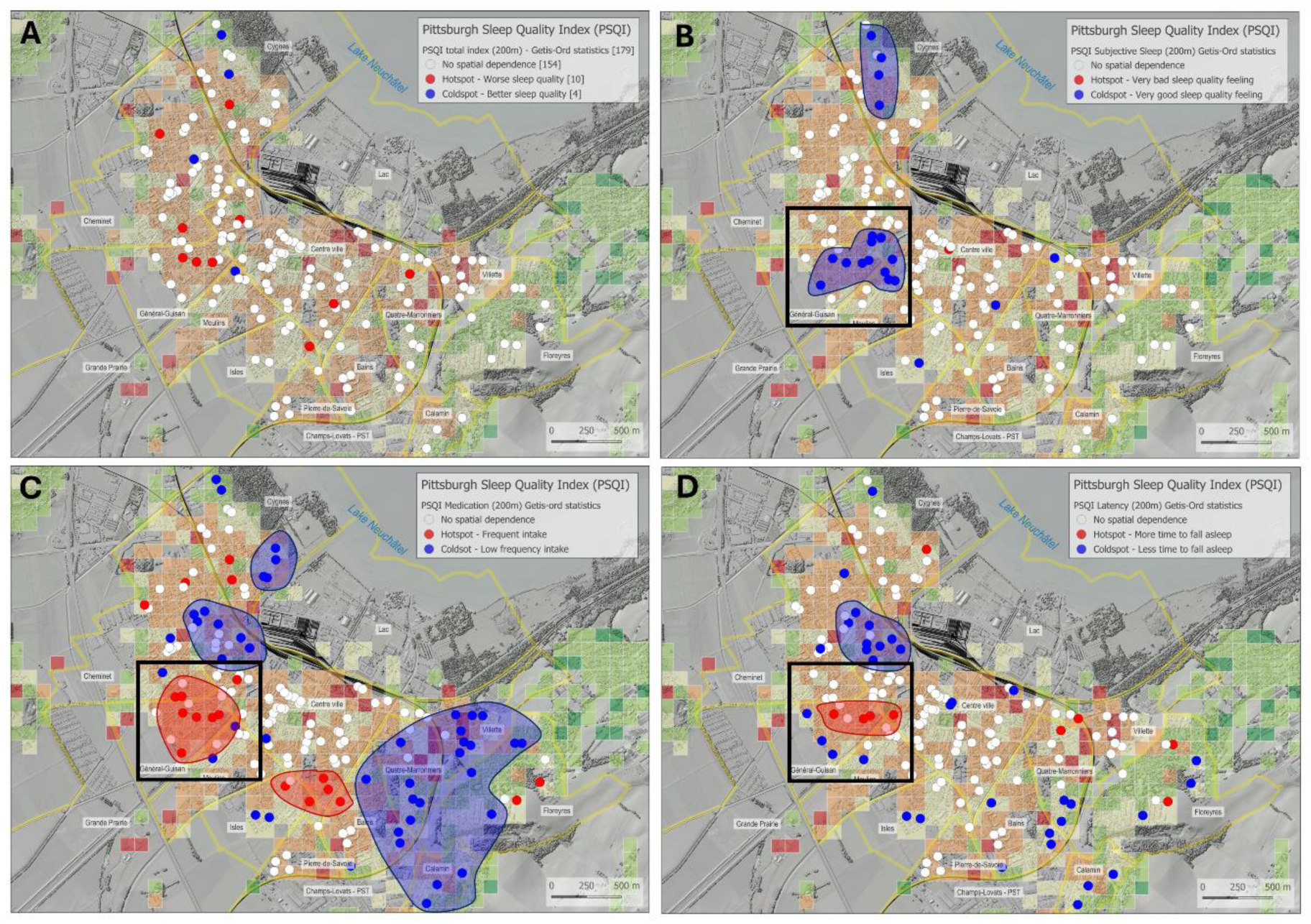
Results of the Getis-Ord analysis for PSQI total score [A] and significant PSQI sub-scores (Subjective Sleep [B]; Medication [C]; Latency [D]). In blue are coldspots revealing a significant below-average pattern of results, and in red hotspots revealing a significant above-average pattern of results. The circled areas highlight clusters of results. Inset in black shows the same neighborhood of the town of Yverdon-les-Bains, with different patterns of results according to the PSQI sub-scores, revealing better subjectively perceived sleep, but associated with higher medication intake and poorer sleep latency. Moreover, in the background a thematic map of the city of Yverdon-les-Bains showing inhabited hectares and the value of the Eurostat At Risk of Poverty (AROP) indicator [21]. This relative poverty indicator uses a cut-off value corresponding to 60% of the yearly median income of the administrative unit of reference (here the canton of Vaud for 2022). Hectares in dark red show a median income under the poverty threshold. In orange are shown the hectares just above this threshold. Green and dark green hectares show wealthy neighborhoods grouped in the eastern part of the city, while places where people receive an income just above the precariousness threshold can be found throughout the rest of this urban area. Deprived zones are scattered toward the suburbs of Yverdon-les-Bains. The background map is the swissSURFACE3D layer produced by ©swisstopo (https://www.swisstopo.admin.ch/fr/modele-altimetrique-swisssurface3d).

## 4. Discussion

The aim of this study was to assess the implementation of the Urbasan project in the town of Yverdon-les-Bains, focusing on physical and mental health while considering environmental factors. The study was conducted on a representative sample and sought to explore the relationship between health outcomes and environmental variables. The analyses revealed a high prevalence of sleep disorders and daytime drowsiness among residents. Additionally, significant negative associations were found between sleep scores and several environmental variables, including Land Surface Temperature (LST), Normalized Difference Vegetation Index (NDVI), Atmospherically Resistant Vegetation Index (ARVI), road/rail noise, and the Risk Of Poverty Index (AROP), as well as a positive a positive association between PSQI – Subjective Sleep and ARVI. Finally, the spatial analysis of the total sleep impairment score, which showed a high overall level of impairment in the behavioral results, did not reveal any specific spatial clusters of general sleep impairment, likely due to the high mean score across the entire population. However, the analysis of the PSQI subscores revealed distinct spatial patterns, suggesting interactions between different sleep factors and their impairment across specific spatial areas within the city.

First, from a behavioral and epidemiological perspective, our results reveal a high prevalence of self-reported sleep disorders within the general population, highlighting the importance of systematically assessing sleep-related symptoms and understanding how they may be influenced by the urban environment. The prevalence observed for sleep quality in our study was higher than that reported in other Swiss cities (e.g., in Geneva approximately 50% of a representative sample had pathological PSQI scores, [26]), while the prevalence of daytime sleepiness was comparable to that observed in medium sized cities in Canada [27]. This discrepancy for sleep quality may be explained by the greater impact of environmental variables specific to Yverdon-les-Bains, particularly in certain neighborhoods. These findings confirm the need for local epidemiological studies, which could uncover disparities even within neighboring cities in the same country. This also underscores the importance of widespread sleep assessments and early interventions, ranging from short- to long-term strategies within urban planning, to prevent the chronicization of these disorders and mitigate their potential long-term negative impacts, both clinically and socioeconomically.

Second, our analysis revealed significant negative associations between subjective sleep quality (as measured by PSQI subscores) and medication use with various environmental variables. These findings suggest a complex interplay of environmental and socio-economic factors contributing to sleep disturbances [1]. While previous studies have highlighted the relationships between these variables, often in isolation, our study uniquely associates them with specific geographic clusters within the city, providing new insights into localized patterns of sleep impairment. Our findings underscore the value of collaborative cohort studies in evaluating sleep disorders [7], offering a foundation for targeted and precise interventions by public authorities, specifically, on the basis of the PSQI sub-scores which make it possible to isolate specific factors of sleep disorders at specific spatial clusters, revealing the need for a precision approach in the context of mitigation intervention. Such interventions could help mitigate the environmental factors influencing sleep quality, particularly in certain districts of the city.

Third, and finally, in an exploratory manner, we associated sleep data with geospatial data, marking, to the best of our knowledge, the first attempt not only to consider sleep disorders in their entirety but also to evaluate their sub-factors, such as the PSQI subscores. This approach allowed us to identify that, within a context of widespread sleep disturbances, specific clusters of results emerge. These findings suggest that, spatially, the factors influencing sleep disturbances are not uniform. Specifically, individuals in certain spatial clusters may experience sleep problems linked to interactions between sleep sub-factors. For example, while some subjective clusters report better sleep than the average, these same clusters tend to take more medication and have longer sleep latency, revealing a discrepancy between subjective sleep perception and objective sleep measures. Thus, these results provide insight into how interventions can be precisely tailored to specific regions or neighborhoods within a city, potentially mitigating sleep disturbances on a broader scale.

Nevertheless, our study suffers from certain limitations. Firstly, our sample may not be entirely representative, and the reproducibility of the results may be limited, despite the large number of participants as compared to previous studies. The Urbasan project aims to recruit 2,000 participants, thus allowing the results to be re-evaluated in the future. Secondly, we are using self-questionnaires, which are a subjective assessment of sleep quality, which is not equivalent to an objective assessment. Thirdly, these data are from a specific city and may not be reproducible in other contexts, nevertheless, the associated method provides an approach that is adapted and validated in other contexts. One promising way to further improve the granularity and precision of the outcomes of such collaborative studies such as ours would be to implement measuring in a representative sub-cohort sleep with over-night monitoring and using wearables providing data such as exposure to pollutants, 24h heart rate and accelerometry.

## 5. Conclusions

The Urbasan collaborative project proves to be a valuable tool for evaluating health outcomes in relation to environmental using the georeferenced places of residence of the subjects. In this context, sleep disorders are highly prevalent and should be assessed within the framework of daily life, considering the associated environmental factors. This approach allows for the identification of geographic clusters of sleep disturbances, thereby enabling more precise, targeted interventions. Collaborative projects like Urbasan are essential to better depict regional specificities and guide public health policies to address local needs effectively in order to improve population health.

## Acknowledgement

We thank Dimitrios Lampropoulos, Raphaël Bize, Mattia Vacchiano, Camille Ducrey Poroes, Albert Gaspoz, Grégory Dozot, and Robin Zweifel for their contribution to the group discussions, to the elaboration of communication medias, or for their work related to the running and the maintenance of the Urbasan website.

